# Evaluating the impact of COVID-19 on the HIV epidemic among men who have sex with men in Australia: A modelling study

**DOI:** 10.1101/2024.12.15.24318054

**Authors:** Rongxing Weng, Jisoo A. Kwon, Mo Hammoud, Brent Clifton, Nick Scott, Skye McGregor, Richard T. Gray

## Abstract

**Introduction:** The initial phase of the coronavirus disease 2019 (COVID-19) pandemic caused significant interruptions to human immunodeficiency virus (HIV) prevention and healthcare services. In Australia, these interruptions coincided with a large reduction in HIV diagnoses but it is unknown if this reflects a reduction in incidence or barriers to testing. We aimed to understand the changes in HIV transmission among men who have sex with men in Australia and the impact of disruptions to HIV prevention, testing and treatment services due to the COVID-19 pandemic and the public health response by incorporating time-sensitive factors in real-world situations.

**Methods:** We developed a mathematical model to estimate monthly HIV incidence between January 2020 and August 2022. We obtained aggregated monthly data for sexual partners, condom use, HIV testing, pre-exposure prophylaxis (PrEP) use, and migration. Three scenarios were simulated: 1) a COVID scenario with all changes in place; 2) a no COVID-19 scenario where input parameters remained at pre-COVID-19 values, and 3) a no COVID-19 scenario with continued PrEP scale-up.

**Results:** In the absence of the COVID-19 pandemic, the estimated number of cumulative infections from January 2020 to August 2022 would have been 1,266 [95% Percentile Interval (PI): 1,100–1,466] compared to 915 [95% PI: 729–1,181] for the COVID-19 scenario (a 27.7% reduction). The largest reduction in infections (44.3%) occurred in 2020 with 273 (95% PI: 221– 344) infections versus 490 (95% PI: 435–552) in the no-COVID scenario. There was a rebound with 387 infections (95% PI: 307–502) in 2021, followed by a reduction to a stable level by August 2022. Our model identified reductions in sexual partners as the leading factor contributing to the change in HIV infections and diagnoses (−24.8% and −10.5%, respectively).

**Conclusions:** A substantial reduction in new HIV infections and diagnoses in Australia occurred during the early stages of the COVID-19 pandemic, which was largely due to reduced HIV transmission. A rebound in infections as sexual partnerships increased between 2021-2022, highlights the imperative to maintain vigorous response efforts and take advantage of the gains made to virtually eliminate HIV transmission in Australia.

## Introduction

The coronavirus disease 2019 (COVID-19) not only resulted in millions of confirmed cases and deaths worldwide [1] but also severely interrupted healthcare use [2]. This led to fewer clinic visits, admissions, tests and treatments [2], potentially increasing the burden of infectious diseases including human immunodeficiency virus (HIV), tuberculosis, and malaria [3].

In the early stages of the COVID-19 pandemic, Australian state and federal governments introduced border closure and lockdown measures to control the spread of COVID-19 [4, 5]. Physical restrictions including physical-distancing rules, stay-at-home orders, and closure of non-essential businesses were also introduced [4]. The COVID-19 vaccine rollout in Australia was launched in late February 2021 [6]. Most restrictions were lifted in December 2021 when more than 90% of people over the age of 16 were fully vaccinated [7], and the international borders were reopened to all fully vaccinated visa holders in February 2022 [8]. These measures significantly influenced sexual behaviours and the use of HIV services [4, 5]. Studies reported a substantial reduction in HIV tests during lockdowns and a dramatic decrease in sexual contacts during the first month of the national lockdown [4, 5]. These changes likely contributed to the large reduction (40%) in the number of HIV diagnoses in Australia, decreasing from 895 in 2019 to 541 in 2021 [9]. However, it is unknown if this reduction in HIV diagnoses is mainly related to a fall in testing due to the decreased access to HIV services [4] or a fall in new infections due to reduced sexual contacts [5, 10]. Another factor could have been border closures and the reduced number of migrant arrivals from overseas. Furthermore, the pandemic affected the use of preventive measures such as pre-exposure prophylaxis (PrEP) and condoms. A previous study in Australia showed that a large proportion (42%) of men who have sex with men (MSM) discontinued PrEP use in 2020 [11]. Similar impacts were reported in other high-income settings with sexual minority men in the United States reporting decreased condomless anal sex with casual partners during the early stages of the COVID-19 pandemic [12]. However, the subsequent influence of these changes on the HIV epidemic remains unclear. To better understand the impacts of the COVID-19 pandemic on the HIV epidemic, an evaluation incorporating all factors related to HIV transmission affected by COVID-19 is required.

The impacts of COVID-19 on the HIV epidemic [13–16] have been assessed through modelling. However, many studies were published in the early stage of the pandemic and relied on assumed impacts on factors related to HIV transmission, rather than actual directly observed impacts. Most modelling studies assumed constant impacts and often overlooked the dynamic impacts of specific policies such as lockdowns and border closures on HIV transmission [13–16]. This highlights the need for more nuanced studies that examine the dynamics of the HIV epidemic in response to the COVID-19 response.

MSM are one of the key populations affected by HIV [17]. In Australia, male-to-male sex is the primary HIV risk exposure, accounting for 70% of new HIV infections between 2015 and 2019 [9] and most HIV infections among MSM are attributable to sex with non-romantic partners (casual sexual partners and friends with benefits) [18]. Detailed datasets are available for this population in Australia. This study aimed to use a mathematical model to evaluate the impact of COVID-19 restrictions on the HIV epidemic among MSM in Australia, considering the empirically recorded monthly changes in sexual behaviours, HIV testing, and migration during the acute phase of the COVID-19 pandemic.

## Methods

We first developed a simple but general and flexible HIV transmission model which we then applied to the MSM population in Australia. This Simple HIV Model is written in R version 4.3.2. and is publicly available under an open access license [19]. No ethical approval or consent was required as the study only used publicly available data.

### Simple HIV Model

Our Simple HIV Model expands previously developed models [20–22]. The model uses difference equations to describe the HIV cascade and the effect of changes in sexual partners, condom use, HIV testing, and PrEP use on HIV transmission. At a pre-specified timestep *t*, *N*_*t*_ is the number of people living with HIV in the population, *I*_*t*_ is the number of new infections, and *β*_*t*_ is the transmission probability from a person with an unsuppressed HIV viral load to a susceptible partner not on PrEP. Therefore, if no interventions were implemented, HIV-positive individuals would transmit *N*_*t*_*β*_*t*_ infections at timestep *t*. To represent the steps of the HIV cascade, we defined *d*_*t*_ to be the proportion of people living with HIV who are diagnosed, *τ*_*t*_ to be the proportion of those diagnosed on antiretroviral therapy (ART), *σ*_*t*_ to be the proportion of those on ART with suppressed viral load (< 200 copies per ml). If *ϕ* is the reduction in transmission due to viral suppression then HIV-positive individuals transmit *N*_*t*_*β*_*t*_(1 − *d*_*t*_*τ*_*t*_*σ*_*t*_*ϕ*) infections at timestep *t*. To account for PrEP use, we defined *ω*_*t*_ to be the proportion of the susceptible population on PrEP, and *∊*_*p*_ to be the efficacy of PrEP in preventing infection. The number of infections at time step t then becomes *N*_*t*_*β*_*t*_(1 − *d*_*t*_*τ*_*t*_*σ*_*t*_*ϕ*)(1 − *ω*_*t*_*∊*_*p*_). With these definitions, a simple risk equation was constructed to estimate the number of new infections due to transmission to susceptible sexual partners:

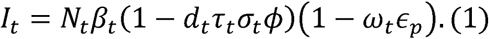

To account for changes in the population due to migration and mortality, we defined *M*_*t*_ to be the number of people who enter Australia living with HIV, *E*_*t*_ to be the number of people who leave Australia living with HIV, and *μ*_*t*_ to be the probability of all-cause mortality in people living with HIV. Then the number of people living with HIV in the population at timestep *t*+1 would be (*N*_*t*_ + *I*_*t*_ + *M*_*t*_ − *E*_*t*_)(1 − *μ*_*t*_). These definitions allowed us to construct Equation 2:

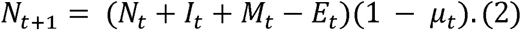

Before calculating *N*_*t*_ and *I*_*t*_, an initial value for the transmission probability (*β*_0_) at time t = 0 is required. This is obtained using initial values for the population size, new infections, and each parameter in the following equation:

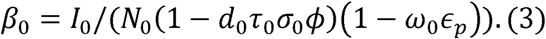

### Effect of changing sexual behaviours

To consider the impacts of changes in sexual behaviours, we incorporated the effects of changes in condom use and sexual partners. If the proportion of condom use changes to *c*_*t*_ at timestep *t* compared to an initial level *c*_0_ with an effectiveness of *∊*_*c*_, the relative change in the transmission probability is given by 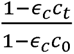. If the number of sexual partners changes to *p_t_* of the time compared to a pre-covid level of *p*_0_, the relative change in the transmission probability is given by 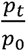.

By applying these relative changes to the initial transmission probability, we obtain:

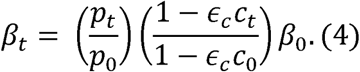

### Effect of changes in testing rates

The proportion diagnosed over time is affected by changes in testing rates. If the proportion of the population tested at time-step t is *T*_*t*_, the number of people living with HIV diagnosed at the next time step is given by:

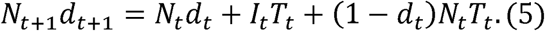

With Equation 2, this can be used to calculate the proportion diagnosed at the next time step:

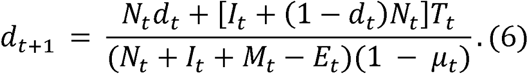

We only included diagnosed immigrants (*M*_*t*_) and emigrants (*E*_*t*_) in the model assuming the number undiagnosed is small compared to the overall population and has a minimal impact on the results.

Defining the number of new diagnoses at the initial timestep to be *D*_0_, we solved Equation 7 for the proportion of the population tested at the initial timestep (*T*_0_) before calculating *d*_*t*+1_:

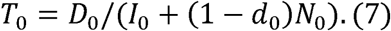

### Application of model

For this study, we focused on the MSM population in Australia and used a monthly timestep starting from December 2019. We used notifications from national surveillance as a proxy for the initial number of diagnosed cases (*D*_0_) [9]. The changes in each parameter were captured over time following the implementation of restrictions and border closures in response to the COVID-19 outbreak in March/April 2020 [23]. Up to that time we assumed input parameters remained at their December 2019 values. To estimate the impact of the COVID-19 pandemic on sexual partners, we used the reported number of non-romantic partners (casual sexual partners and friends with benefits) as a proxy for the number of sexual partners at both time 0 (*p*_0_) and time *t* (*p*_*t*_). To estimate the impact of the COVID-19 pandemic on condom use, we used the reported proportion of MSM with casual partners reporting no condomless anal intercourse in the past four weeks as a proxy for the proportion of MSM reporting condom use at both time 0 (*c*_0_) and time *t* (*c*_*t*_). Constant parameters are presented in Table 1. The input variables *ω*_*t*_, *T*_*t*_, *c*_*t*_, and *p*_*t*_ are time varying and were estimated using data from Kirby Institute’s monitoring of PrEP uptake in Australia, the Flux cohort study, and the Gay Community Periodic Survey [5, 24–26] (details provided in the Supplementary Material).

**Table 1.**
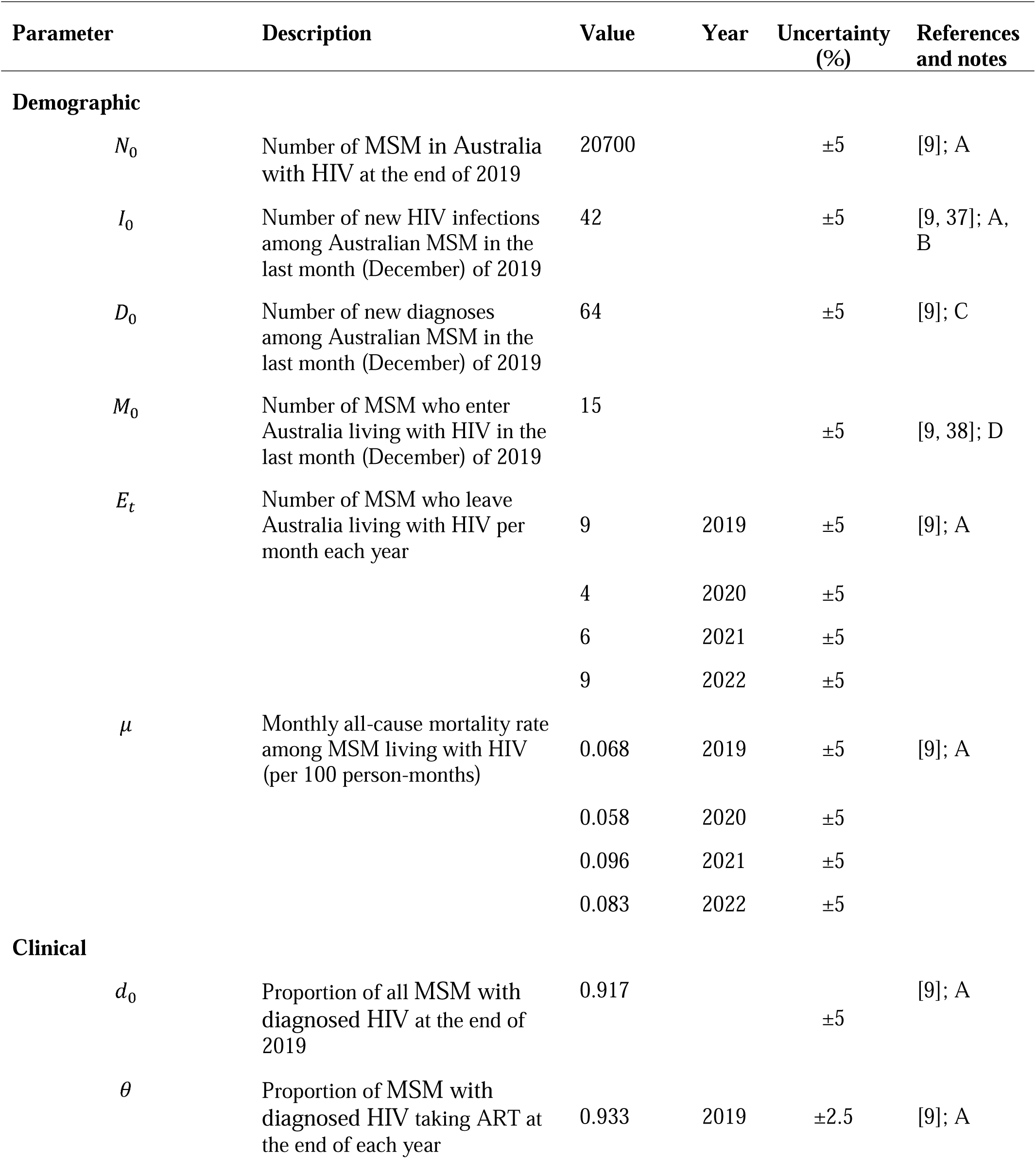
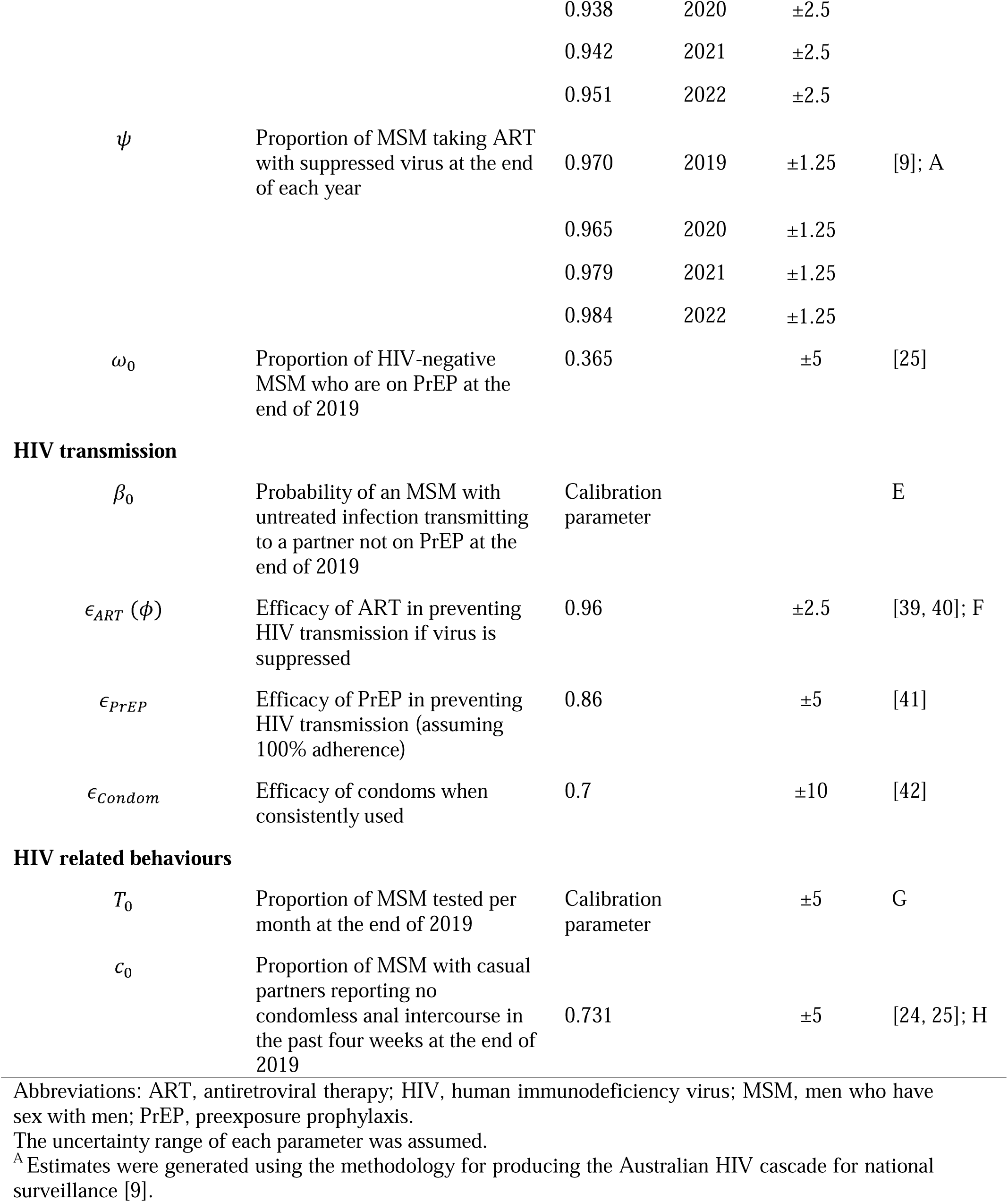
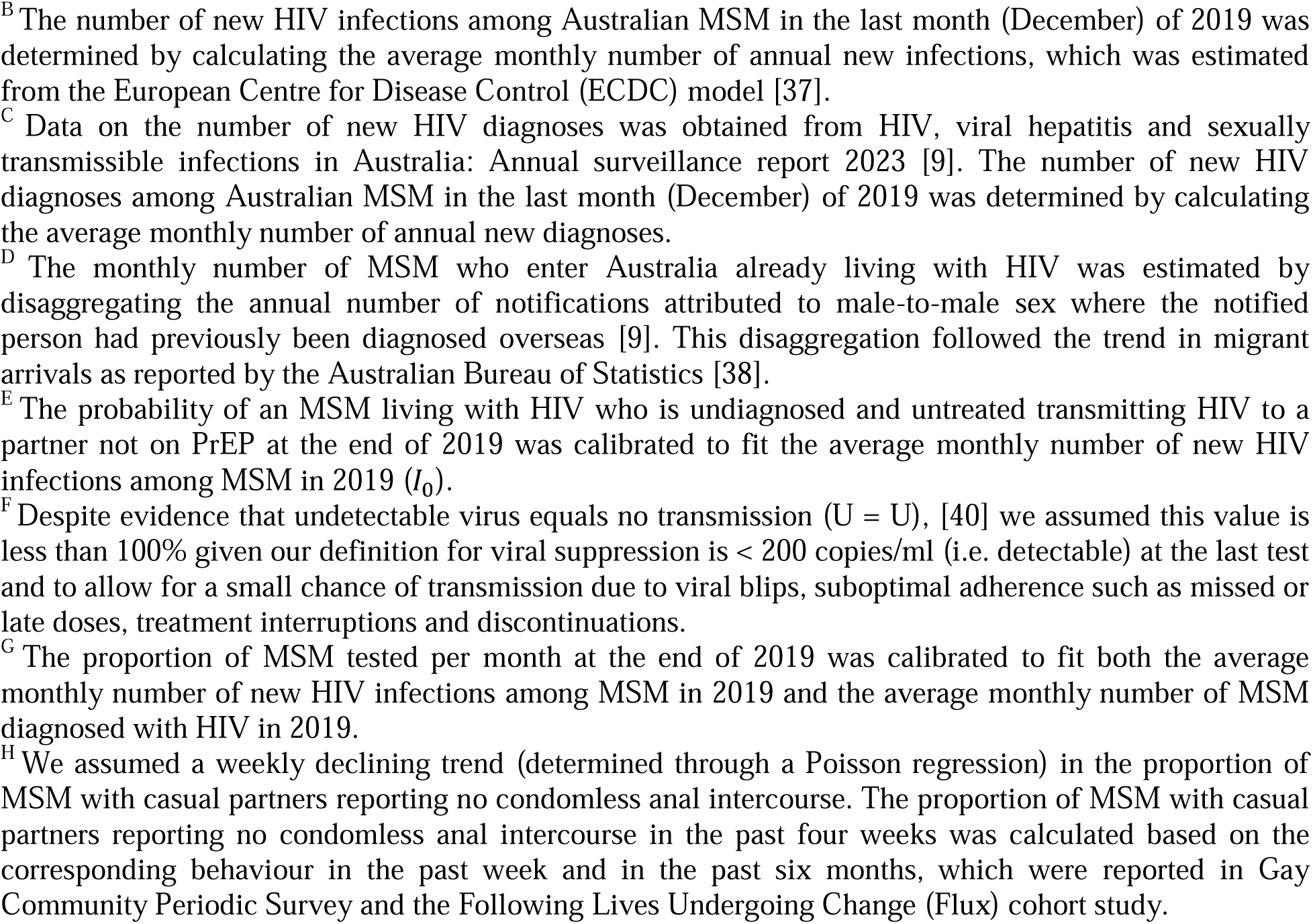
Constant input parameters and their associated descriptions and values (with uncertainty bounds) used in the model.

| Parameter | Description | Value | Year | Uncertainty (%) | References and notes |
| --- | --- | --- | --- | --- | --- |
| Demographic |  |  |  |  |  |
| $N_0$ | Number of MSM in Australia with HIV at the end of 2019 | 20700 | | $\pm 5$ | [9]; A |
| $I_0$ | Number of new HIV infections among Australian MSM in the last month (December) of 2019 | 42 | | $\pm 5$ | [9, 37]; A, B |
| $D_0$ | Number of new diagnoses among Australian MSM in the last month (December) of 2019 | 64 | | $\pm 5$ | [9]; C |
| $M_0$ | Number of MSM who enter Australia living with HIV in the last month (December) of 2019 | 15 | | $\pm 5$ | [9, 38]; D |
| $E_t$ | Number of MSM who leave Australia living with HIV per month each year | 9 | 2019 | $\pm 5$ | [9]; A |
| | | 4 | 2020 | $\pm 5$ | |
| | | 6 | 2021 | $\pm 5$ | |
| | | 9 | 2022 | $\pm 5$ | |
| $\mu$ | Monthly all-cause mortality rate among MSM living with HIV (per 100 person-months) | 0.068 | 2019 | $\pm 5$ | [9]; A |
| | | 0.058 | 2020 | $\pm 5$ | |
| | | 0.096 | 2021 | $\pm 5$ | |
| | | 0.083 | 2022 | $\pm 5$ | |
| Clinical |  |  |  |  |  |
| $d_0$ | Proportion of all MSM with diagnosed HIV at the end of 2019 | 0.917 | | $\pm 5$ | [9]; A |
| $\theta$ | Proportion of MSM with diagnosed HIV taking ART at the end of each year | 0.933 | 2019 | $\pm 2.5$ | [9]; A |
| | | 0.938 | 2020 | $\pm 2.5$ | |
| | | 0.942 | 2021 | $\pm 2.5$ | |
| | | 0.951 | 2022 | $\pm 2.5$ | |
| $\psi$ | Proportion of MSM taking ART with suppressed virus at the end of each year | 0.970 | 2019 | $\pm 1.25$ | [9]; A |
| | | 0.965 | 2020 | $\pm 1.25$ | |
| | | 0.979 | 2021 | $\pm 1.25$ | |
| | | 0.984 | 2022 | $\pm 1.25$ | |
| $\omega_0$ | Proportion of HIV-negative MSM who are on PrEP at the end of 2019 | 0.365 | | $\pm 5$ | [25] |
| **HIV transmission** |  |  |  |  |  |
| $\beta_0$ | Probability of an MSM with untreated infection transmitting to a partner not on PrEP at the end of 2019 | Calibration parameter | | | E |
| $\epsilon_{ART}(\phi)$ | Efficacy of ART in preventing HIV transmission if virus is suppressed | 0.96 | | $\pm 2.5$ | [39, 40]; F |
| $\epsilon_{PrEP}$ | Efficacy of PrEP in preventing HIV transmission (assuming 100% adherence) | 0.86 | | $\pm 5$ | [41] |
| $\epsilon_{Condom}$ | Efficacy of condoms when consistently used | 0.7 | | $\pm 10$ | [42] |
| **HIV related behaviours** |  |  |  |  |  |
| $T_0$ | Proportion of MSM tested per month at the end of 2019 | Calibration parameter | | $\pm 5$ | G |
| $c_0$ | Proportion of MSM with casual partners reporting no condomless anal intercourse in the past four weeks at the end of 2019 | 0.731 | | $\pm 5$ | [24, 25]; H |

Abbreviations: ART, antiretroviral therapy; HIV, human immunodeficiency virus; MSM, men who have sex with men; PrEP, preexposure prophylaxis.
The uncertainty range of each parameter was assumed.
<sup>A</sup> Estimates were generated using the methodology for producing the Australian HIV cascade for national surveillance [9].
<sup>B</sup> The number of new HIV infections among Australian MSM in the last month (December) of 2019 was determined by calculating the average monthly number of annual new infections, which was estimated from the European Centre for Disease Control (ECDC) model [37].
<sup>C</sup> Data on the number of new HIV diagnoses was obtained from HIV, viral hepatitis and sexually transmissible infections in Australia: Annual surveillance report 2023 [9]. The number of new HIV diagnoses among Australian MSM in the last month (December) of 2019 was determined by calculating the average monthly number of annual new diagnoses.
<sup>D</sup> The monthly number of MSM who enter Australia already living with HIV was estimated by disaggregating the annual number of notifications attributed to male-to-male sex where the notified person had previously been diagnosed overseas [9]. This disaggregation followed the trend in migrant arrivals as reported by the Australian Bureau of Statistics [38].
<sup>E</sup> The probability of an MSM living with HIV who is undiagnosed and untreated transmitting HIV to a partner not on PrEP at the end of 2019 was calibrated to fit the average monthly number of new HIV infections among MSM in 2019 ( $I_0$ ).
<sup>F</sup> Despite evidence that undetectable virus equals no transmission ( $U = U$ ), [40] we assumed this value is less than 100% given our definition for viral suppression is $< 200$ copies/ml (i.e. detectable) at the last test and to allow for a small chance of transmission due to viral blips, suboptimal adherence such as missed or late doses, treatment interruptions and discontinuations.
<sup>G</sup> The proportion of MSM tested per month at the end of 2019 was calibrated to fit both the average monthly number of new HIV infections among MSM in 2019 and the average monthly number of MSM diagnosed with HIV in 2019.
<sup>H</sup> We assumed a weekly declining trend (determined through a Poisson regression) in the proportion of MSM with casual partners reporting no condomless anal intercourse. The proportion of MSM with casual partners reporting no condomless anal intercourse in the past four weeks was calculated based on the corresponding behaviour in the past week and in the past six months, which were reported in Gay Community Periodic Survey and the Following Lives Undergoing Change (Flux) cohort study.

### Scenarios

We simulated three scenarios from December 2019 to August 2022. The first scenario was the “COVID-19 scenario”, where all changes in monthly parameter values occur and reflect the impact of COVID-19. The second scenario was the “no COVID-19 scenario”, a counterfactual scenario where the input parameters remained at their values in December 2019 and corresponded to pre-COVID-19 values. The third scenario was the “no COVID-19 plus PrEP scenario”, an alternative counterfactual scenario where PrEP scale-up and the corresponding decrease in condom use prior to 2019 continued during 2020-2022, with the other parameters remaining at their December 2019 value. In this scenario, we assumed that the proportion of MSM on PrEP increased from 36.5% to 58.2% over this period following trends for the number of people dispensed PrEP [26] and the proportion using condoms from 73.1% to 64.9% following the trends reported in the Gay Community Periodic Survey [25] (Table S1). Further details of each scenario are provided in the Supplementary Material.

Further simulations were conducted by separately modelling the impact of COVID-19 on each time series variable to assess their individual effects on the HIV epidemic. Migration was considered as a single variable incorporating both MSM living with HIV who enter (*M*_*t*_) and leave (*E*_*t*_) Australia.

### Model calibration, validation, and uncertainty analysis

We calibrated the transmission probability at the initial timestep (*β*_0_) to fit the average monthly number of new HIV infections among MSM in 2019 using Equation 3 [9]. We then calibrated the testing rate at the initial timestep (*T*_0_) to fit both the average monthly number of new HIV infections among MSM in 2019 and the average monthly number of MSM diagnosed with HIV in 2019 using Equation 7 [9]. The annual numbers of MSM diagnosed with HIV from 2020 to 2022 were then used to validate the results of the model. To account for uncertainty, all three scenarios were simulated 1000 times by randomly sampling from each parameter range. We assumed that the uncertainty range for parameters including PrEP use (*ω*_*t*_), sexual partners (*p*_*t*_), condom use (*c*_*t*_), and testing rate (*T*_*t*_) at each timestep were ±5%. The results from each simulation were then used to derive a 95% percentile interval (95% PI) for the model outputs.

## Results

After calibration, the estimated number of annual diagnoses in the COVID-19 scenario closely aligned with data from national surveillance between 2020-2022 (Figure 1), providing a validation for our model estimates.

**Figure 1.**
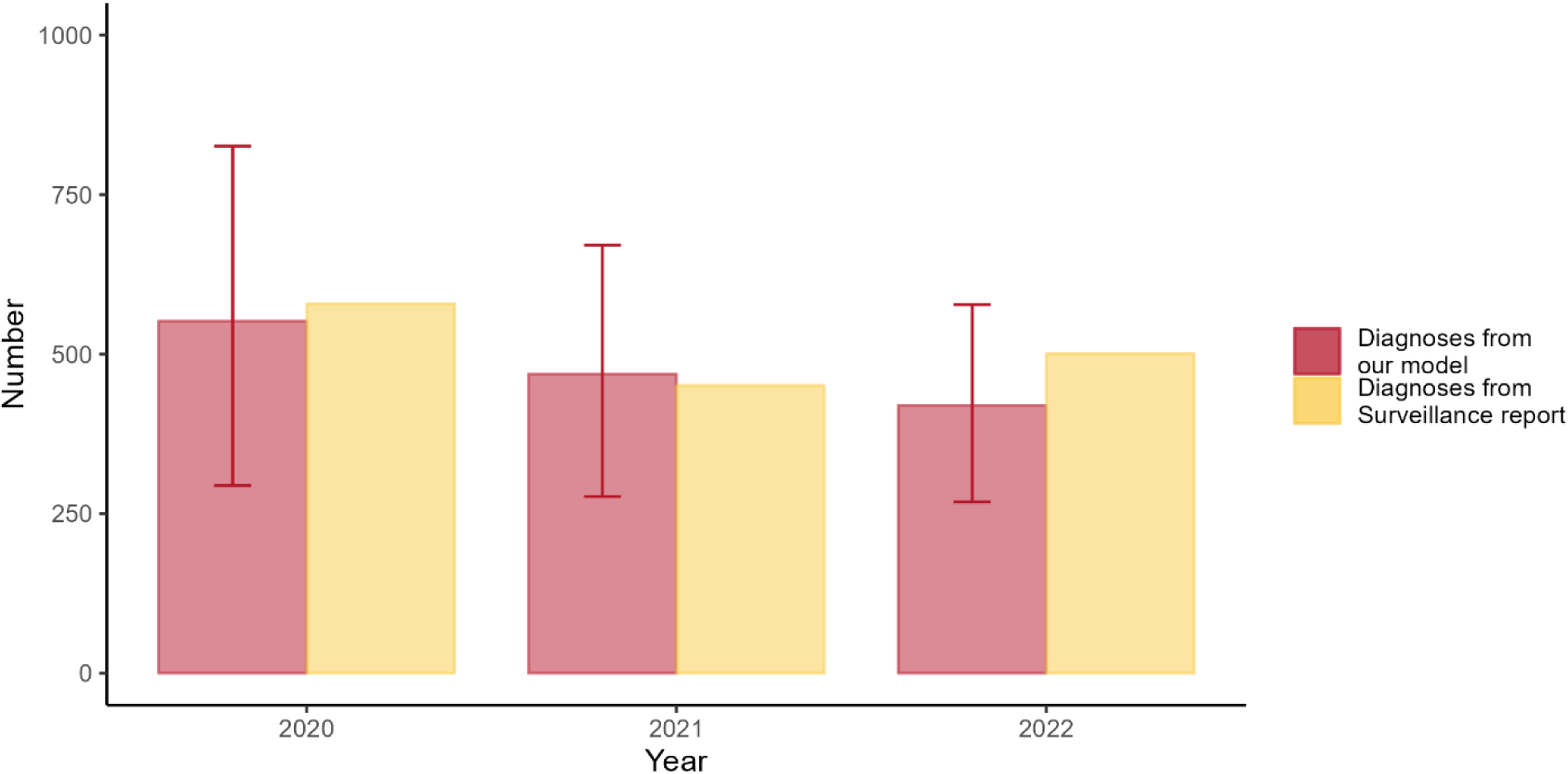
Comparison of estimated diagnoses from the model with diagnoses from national surveillance data. The estimated diagnoses from the model in 2022 were adjusted to reflect the annual number.

### Number of new infections, diagnoses, and MSM with HIV in each scenario

As shown in Table 2 and Figure 2, in the absence of the COVID-19 pandemic (no COVID-19 scenario), the estimated number of cumulative infections from 2020 to 2022 would have been the highest, at 1,266 [95% PI: 1,100–1,466]. Figure 2 shows that there was a stable level of monthly new infections with a slightly decreasing trend. In contrast, there was a 27.7% reduction in the number of cumulative infections for the COVID-19 scenario (915, [95% PI: 729–1,181]) (Table 2). The largest reduction in infections (44.3%) occurred in 2020 with 273 (95% PI: 221–344) infections versus 490 (95% PI: 435–552) for the no COVID-19 scenario. There was a rebound with 387 infections (95% PI: 307–502) in 2021, coinciding with a rebound in sexual partners (see Supplementary Material Figure S1), followed by a reduction to a stable level by August 2022 (Figure 2). An increase in PrEP use (no COVID-19 plus PrEP scenario) would have resulted in a slight reduction in infections compared to the no COVID-19 scenario, but not as substantial as observed in the COVID-19 scenario. The estimated number of cumulative infections from 2020 to 2022 in the no COVID-19 plus PrEP scenario was 1,151 (95% PI: 998–1,339), with a 9.1% reduction compared to the no COVID-19 scenario (Table 2). The monthly difference in the number of new infections between the no COVID-19 scenario and the no COVID-19 plus PrEP scenario expanded over time, with the largest difference in infections (21.1%) occurring in August 2022 (Figure 2). The uncertainty ranges for the three scenarios started to overlap in April 2021.

**Figure 2.**
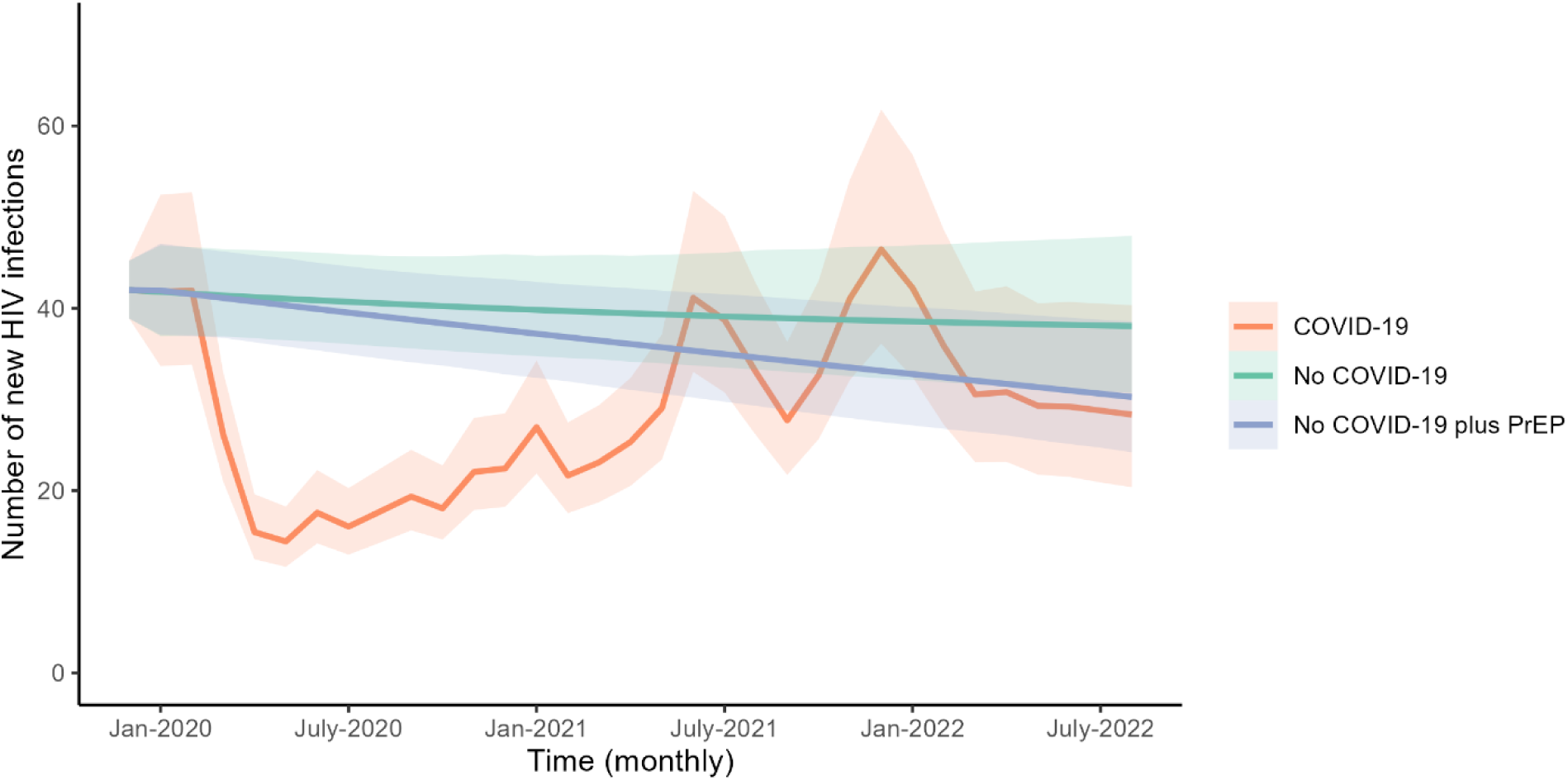
Monthly new HIV infections among men who have sex with men in Australia for each scenario. Bands indicate 95% percentile intervals.

**Table 2.** Number of cumulative infections, cumulative diagnoses, and MSM living with HIV in each scenario between January 2020 to August 2022.

| Variables | Value | Lower bound | Upper bound |
| --- | --- | --- | --- |
| Cumulative infections |  |  |  |
| COVID-19 | 915 | 729 | 1,181 |
| No COVID-19 | 1,266 | 1,100 | 1,466 |
| No COVID-19 plus PrEP | 1,151 | 998 | 1,339 |
| Cumulative diagnoses |  |  |  |
| COVID-19 | 1,301 | 758 | 1,870 |
| No COVID-19 | 1,613 | 1,041 | 2,221 |
| No COVID-19 plus PrEP | 1,579 | 1,006 | 2,189 |
| MSM living with HIV |  |  |  |
| COVID-19 | 21,273 | 20,285 | 22,281 |
| No COVID-19 | 21,724 | 20,757 | 22,725 |
| No COVID-19 plus PrEP | 21,617 | 20,664 | 22,623 |

Abbreviations: HIV, human immunodeficiency virus; MSM, men who have sex with men; PrEP, preexposure prophylaxis.

Similarly, in the no COVID-19 scenario, the number of cumulative diagnoses over the simulation period would have been 1,613 (95% PI: 1,041–2,221) compared to 1,301 (95% PI: 758–1,870) for the COVID-19 scenario (a 19.3% reduction). The projected number of cumulative diagnoses in no COVID-19 plus PrEP scenario was intermediate to the estimates of the other two scenarios, at 1,579 (95% PI: 1,006–2,189) (Table 2). The number of MSM with HIV would have been slightly higher in the no COVID-19 scenario than that in the COVID-19 scenario, increasing from 21,273 (95% PI: 20,285–22,281) to 21,724 (95% PI: 20,757–22,725) by August 2022 (Table 2). Whereas in the no COVID-19 plus PrEP scenario the number of MSM with HIV was intermediate to the estimates of the other two scenarios, at 21,617 (95% PI: 20,664–22,623) (Table 2). Further details are provided in the Supplementary Material.

### Impacts of each time series variable on the HIV epidemic

Our model identified sexual partners as the biggest driver of the changes in new HIV infections (Table S3, Figure S2), with a reduction of 24.8% over the simulation period compared to the no COVID-19 scenario. The other individual variables (HIV testing, condom use, PrEP use, and migration) had smaller effects on infections (2.5% increase, 1.7% reduction, 2.5% increase, and 0.1% reduction, respectively). In terms of HIV diagnoses, the substantial decrease was primarily due to a combination of the reduction in sexual partners (10.5% reduction) and HIV testing (7.5% reduction) over the study period. Further details are provided in the Supplementary Material.

## Discussion

Our study used a mathematical model to evaluate the impact of COVID-19 on the HIV epidemic among MSM in Australia. Our simulations estimated that COVID-19-related interruptions to HIV testing, PrEP use, and migration as well as the associated impact on partner numbers and condom use resulted in a substantial decrease in both diagnoses (19.3%) and new HIV infections (27.7%). The largest reduction in new infections (44.3%) occurred in 2020, followed by a rebound in 2021 and then a reduction to a stable level in 2022, at a similar level to what might happened if COVID-19 did not occur, which suggests that the direct impact of COVID-19 on HIV transmission dynamics may have diminished over time. Our study provides new insights into the impacts of COVID-19 on the HIV epidemic by capturing monthly changes in time-sensitive factors in real-world situations including sexual partners, HIV testing, condom use, PrEP use, and migration, compared to previous studies that relied on assumed data or constant impacts [13–16].

Our study identified reductions in sexual partners, proxied by the number of non-romantic partners, and HIV testing as the primary drivers behind the observed decline in HIV diagnoses. This finding highlights the importance of maintaining HIV testing services when public health restrictions are in place in response to a severe pandemic. Telehealth-delivered, web-based, and home-based HIV testing services could be promoted to ensure continuing HIV testing [27–29]. Also, national surveillance reported an increasing trend in the proportion of late diagnoses among MSM between 2019 and 2022, indicating a COVID-19 lag effect on the HIV epidemic [9].

We also found that physical distancing restrictions and the resulting reduction in sexual partners had the biggest impact on new infections, with a 24.8% decrease alone compared to a 27.7% reduction when all changes were included, counteracting the increased risk of infection due to disrupted HIV testing and PrEP use. Our findings are consistent with previous modelling. A study from the United States found that compared to condom use, PrEP use, and HIV testing, the sexual partner numbers were the biggest driver of the change in new HIV infections and a theoretical 50% decrease in the sexual partner numbers could reduce the number of new HIV infections by 24% [14]. Another modelling study showed that the reduced number of sexual partners could offset the impact of HIV service disruptions on new HIV infections [30]. However, these studies only used theoretical scenarios and did not consider empirically observed monthly changes. Compared to these studies, our study incorporated data on the actual dynamics of these time-sensitive factors in real-world situations.

This study projected that continued PrEP scale-up would have slightly reduced the number of new HIV infections with an intensified impact over time, which is consistent with findings from previous studies [31, 32]. This finding highlights the importance of the continuity and expansion of PrEP use. Despite the COVID-19 pandemic disrupting the increasing trend, PrEP use had fully recovered by October 2021 and has continued to increase steadily until the latest available data in June 2023 [33].

There are several limitations in our study. First, results from the current study should be interpreted with caution due to the simplicity of the model. However, our validation results indicate that our model estimates the number of new HIV diagnoses over the entire simulation period well. Also, the robustness of the time series data for variables such as PrEP use, HIV testing, condom use, and sexual partners supported the reliability of the results from our model. Second, we did not have monthly data on ART use and viral suppression to incorporate into the model. Several studies have highlighted the significant impacts that the changes in ART use and viral suppression during the COVID-19 pandemic can have on the HIV epidemic [14–16]. However, previous studies in Australia showed that the proportion of viral suppression and access to ART essentially continued uninterrupted, even during the lockdown restrictions [34, 35]. This finding suggests that the uncertainty introduced by the lack of monthly data on ART use and viral suppression is likely minimal. Third, our results may not fully capture the impacts of key variables on the HIV epidemic during March and April 2020, as the Flux study only started in May 2020. We assumed that the value of these variables in April 2020 was the same as that in May 2020. Given that restrictions and the border closure were introduced in mid-March 2020, we assumed the impact on these variables for March was half of that in April. Our model may underestimate the impact of restrictions on these variables in March and April 2020, as some restrictions were eased in May 2020 [36]. Fourth, we considered the overall impact of lockdown measures on the HIV epidemic. However, these measures varied significantly between states and territories. These regional differences in public health responses could have had varying impacts on factors related to HIV transmission, potentially influencing our results. Future analyses could explore these jurisdictional variations to better understand the HIV epidemic at the state and territory level during the pandemic. Fifth, potential discrepancies arise because we used the proportion of MSM with casual partners reporting no condomless anal intercourse in the past four weeks as a proxy for condom use and the number of sexual partners to represent overall sexual activity rather than consider sexual acts, as a proxy for sexual activity. Finally, in our model, we assumed that changes in condom use and PrEP use are independent. This assumption may not fully reflect real-world behaviours, as those taking up PrEP are also the ones who are reducing condom use. By applying relative risks independently, the model may overestimate HIV transmission risk and underestimate the impact of PrEP use.

Our study is the first study to incorporate the time varying effects of COVID-19 restrictions on the HIV epidemic among MSM using real-world data. Our study demonstrates that COVID-19 restrictions contributed to a genuine reduction in new HIV infections in Australia. The generalisability and flexibility of the Simple HIV model means it is adaptable to other settings or populations.

## Conclusions

A substantial reduction in new HIV infections and diagnoses in Australia occurred during the early stages of the COVID-19 pandemic, which was largely due to reduced HIV sexual partnerships. A rebound in infections between 2021-2022, highlights the imperative to maintain vigorous response efforts and take advantage of the gains made to virtually eliminate HIV transmission in Australia. This outcome shows how changes in time-sensitive factors in response to large-scale public health policies or interventions can affect HIV epidemics. By understanding this relationship, public health authorities can better prepare for and mitigate the effects of similar disruptions on the HIV epidemic in the future.

## Authors’ contributions

RW and RTG conceived the study. RW and RTG developed the model. RW set up and ran the scenarios. RW drafted the manuscript. JAK, MH, SM, and RTG validated the model input. All authors were involved in writing and revising the manuscript.

## Acknowledgements

We would like to thank Dr Doug Fraser for the information on the PrEP uptake in Australia.

## Data Availability Statement

All data produced in the present work are contained in the manuscript and Supplementary Materials. The Simple HIV Model code used to produce results for this study along with model input parameters and summary results are available online at https://github.com/The-Kirby-Institute/Simple_HIV_model/

## Supporting Information

Supporting Information file 1: Supplementary Material

docx. Additional methodological details and results are provided in the file.

## References

1. Su S, Zhao Y, Zeng N, Liu X, Zheng Y, Sun J, et al. Epidemiology, clinical presentation, pathophysiology, and management of long COVID: an update. Molecular psychiatry. 2023 Oct;28(10):4056–69.

2. Moynihan R, Sanders S, Michaleff ZA, Scott AM, Clark J, To EJ, et al. Impact of COVID-19 pandemic on utilisation of healthcare services: a systematic review. BMJ open. 2021 Mar 16;11(3):e045343.

3. Hogan AB, Jewell BL, Sherrard-Smith E, Vesga JF, Watson OJ, Whittaker C, et al. Potential impact of the COVID-19 pandemic on HIV, tuberculosis, and malaria in low-income and middle-income countries: a modelling study. The Lancet Global health. 2020 Sep;8(9):e1132–e41.

4. Chow EPF, Ong JJ, Donovan B, Foster R, Phillips TR, McNulty A, et al. Comparing HIV Post-Exposure Prophylaxis, Testing, and New Diagnoses in Two Australian Cities with Different Lockdown Measures during the COVID-19 Pandemic. International journal of environmental research and public health. 2021 Oct 14;18(20).

5. Hammoud MA, Maher L, Holt M, Degenhardt L, Jin F, Murphy D, et al. Physical Distancing Due to COVID-19 Disrupts Sexual Behaviors Among Gay and Bisexual Men in Australia: Implications for Trends in HIV and Other Sexually Transmissible Infections. Journal of acquired immune deficiency syndromes (1999). 2020 Nov 1;85(3):309–15.

6. Hanly M, Churches T, Fitzgerald O, MacIntyre CR, Jorm L. Vaccinating Australia: How long will it take? Vaccine. 2022 Apr 14;40(17):2491–7.

7. Khalil S. Covid: Australians desperate for tests amid Omicron surge: BBC; 2022 [cited 2024 23rd June]. Available from: https://www.bbc.com/news/world-australia-59864428.

8. Andrews K. Reopening to tourists and other international travellers to secure our economic recovery: Australian Government; 2022 [updated 07 February 2022; cited 2024 23rd June]. Available from: https://minister.homeaffairs.gov.au/KarenAndrews/Pages/reopening-to-tourists-and-other-international-travellers-to-secure-our-economic-recovery.aspx.

9. King J, McManus H, Kwon A, Gray R, McGregor S. HIV, viral hepatitis and sexually transmissible infections in Australia: Annual surveillance report 2023. The Kirby Institute, UNSW Sydney, Sydney, Australia. 2023.

10. Coombe J, Kong FYS, Bittleston H, Williams H, Tomnay J, Vaisey A, et al. Love during lockdown: findings from an online survey examining the impact of COVID-19 on the sexual health of people living in Australia. Sexually transmitted infections. 2021 Aug;97(5):357–62.

11. Hammoud MA, Grulich A, Holt M, Maher L, Murphy D, Jin F, et al. Substantial Decline in Use of HIV Preexposure Prophylaxis Following Introduction of COVID-19 Physical Distancing Restrictions in Australia: Results From a Prospective Observational Study of Gay and Bisexual Men. Journal of acquired immune deficiency syndromes (1999). 2021 Jan 1;86(1):22–30.

12. Starks TJ, Jones SS, Sauermilch D, Benedict M, Adebayo T, Cain D, et al. Evaluating the impact of COVID-19: A cohort comparison study of drug use and risky sexual behavior among sexual minority men in the U.S.A. Drug and alcohol dependence. 2020 Nov 1;216:108260.

13. Zang X, Krebs E, Chen S, Piske M, Armstrong WS, Behrends CN, et al. The Potential Epidemiological Impact of Coronavirus Disease 2019 (COVID-19) on the Human Immunodeficiency Virus (HIV) Epidemic and the Cost-effectiveness of Linked, Opt-out HIV Testing: A Modeling Study in 6 US Cities. Clinical infectious diseases : an official publication of the Infectious Diseases Society of America. 2021 Jun 1;72(11):e828–e34.

14. Mitchell KM, Dimitrov D, Silhol R, Geidelberg L, Moore M, Liu A, et al. The potential effect of COVID-19-related disruptions on HIV incidence and HIV-related mortality among men who have sex with men in the USA: a modelling study. The lancet HIV. 2021 Apr;8(4):e206–e15.

15. Jewell BL, Smith JA, Hallett TB. Understanding the impact of interruptions to HIV services during the COVID-19 pandemic: A modelling study. EClinicalMedicine. 2020 Sep;26:100483.

16. Jewell BL, Mudimu E, Stover J, Ten Brink D, Phillips AN, Smith JA, et al. Potential effects of disruption to HIV programmes in sub-Saharan Africa caused by COVID-19: results from multiple mathematical models. The lancet HIV. 2020 Sep;7(9):e629–e40.

17. WHO Guidelines Approved by the Guidelines Review Committee. Consolidated Guidelines on HIV Prevention, Diagnosis, Treatment and Care for Key Populations – 2016 Update. Geneva: World Health Organization Copyright © World Health Organization 2016.; 2016.

18. Down I, Ellard J, Bavinton BR, Brown G, Prestage G. In Australia, Most HIV Infections Among Gay and Bisexual Men are Attributable to Sex with ‘New’ Partners. AIDS and behavior. 2017 Aug;21(8):2543–50.

19. Weng R, Gray RT. The-Kirby-Institute/Simple_HIV_model. Zenodo. 10.5281/zenodo.14213772. Updated code available at: https://github.com/The-Kirby-Institute/Simple_HIV_model2024 [

20. Scott N, Stoové M, Kelly SL, Wilson DP, Hellard ME. Achieving 90-90-90 Human Immunodeficiency Virus (HIV) Targets Will Not Be Enough to Achieve the HIV Incidence Reduction Target in Australia. Clinical infectious diseases : an official publication of the Infectious Diseases Society of America. 2018 Mar 19;66(7):1019–23.

21. Kelly SL, Wilson DP. HIV Cascade Monitoring and Simple Modeling Reveal Potential for Reductions in HIV Incidence. Journal of acquired immune deficiency syndromes (1999). 2015 Jul 1;69(3):257–63.

22. Gray RT. Impact of increased antiretroviral therapy use during the treatment as prevention era in Australia. Sexual health. 2023 Jul;20(3):202–10.

23. Cook MJ, Dri GG, Logan P, Tan JB, Flahault A. COVID-19 Down Under: Australia’s Initial Pandemic Experience. International journal of environmental research and public health. 2020 Dec 1;17(23).

24. The Kirby Institute. The Flux Study. UNSW Sydney [cited 2024 May 23th]. Available from: https://www.kirby.unsw.edu.au/research/projects/flux.

25. Chan C, Broady T, Bavinton B, Mao L, Molyneux A, Delhomme F, et al. Gay Community Periodic Survey: Sydney 2022. Sydney: Centre for Social Research in Health, UNSW Sydney; 2022.

26. Fraser D, Medland N, McManus H, Guy R, Grulich A, Bavinton B. Monitoring HIV pre-exposure prophylaxis (PrEP) uptake in Australia (Issue 8). Sydney: The Kirby Institute, UNSW Sydney; 2023.

27. Stephenson R, Sullivan SP, Mitchell JW, Johnson BA, Sullvian PS. Efficacy of a Telehealth Delivered Couples’ HIV Counseling and Testing (CHTC) Intervention to Improve Formation and Adherence to Safer Sexual Agreements Among Male Couples in the US: Results from a Randomized Control Trial. AIDS and behavior. 2022 Aug;26(8):2813–24.

28. Cardwell ET, Ludwick T, Fairley C, Bourne C, Chang S, Hocking JS, et al. Web-Based STI/HIV Testing Services Available for Access in Australia: Systematic Search and Analysis. Journal of medical Internet research. 2023 Sep 22;25:e45695.

29. Zhang Y, Holt M, Chan C, Applegate TL, Bavinton BR, Broady TR, et al. National Surveillance of Home-Based HIV Testing Among Australian Gay and Bisexual Men, 2018-2020: Uptake After Commercial Availability of HIV Self-Tests. AIDS and behavior. 2023 Dec;27(12):4106–13.

30. Jenness SM, Le Guillou A, Chandra C, Mann LM, Sanchez T, Westreich D, et al. Projected HIV and Bacterial Sexually Transmitted Infection Incidence Following COVID-19-Related Sexual Distancing and Clinical Service Interruption. The Journal of infectious diseases. 2021 Mar 29;223(6):1019–28.

31. Stansfield SE, Heitner J, Mitchell KM, Doyle CM, Milwid RM, Moore M, et al. Population-level impact of expanding PrEP coverage by offering long-acting injectable PrEP to MSM in three high-resource settings: a model comparison analysis. Journal of the International AIDS Society. 2023 Jul;26 Suppl 2(Suppl 2):e26109.

32. Doyle CM, Milwid RM, Cox J, Xia Y, Lambert G, Tremblay C, et al. Population-level effectiveness of pre-exposure prophylaxis for HIV prevention among men who have sex with men in Montréal (Canada): a modelling study of surveillance and survey data. Journal of the International AIDS Society. 2023 Dec;26(12):e26194.

33. Fraser D, Medland N, McManus H, Guy R, Grulich A, Chan C, et al. Monitoring HIV pre-exposure prophylaxis (PrEP) uptake in Australia (Issue 9). Sydney: The Kirby Institute, UNSW Sydney; 2023.

34. Lee D, Chow EPF, Aguirre I, Fairley CK, Ong JJ. Access to HIV Antiretroviral Therapy among People Living with HIV in Melbourne during the COVID-19 Pandemic. International journal of environmental research and public health. 2021 Dec 3;18(23).

35. Weerasuria M, Ko C, Ehm A, O’Bryan J, McMahon J, Woolley I, et al. The Impact of the COVID-19 Pandemic on People Living with HIV in Victoria, Australia. AIDS research and human retroviruses. 2021 Apr;37(4):322–8.

36. Storen R, Corrigan NJRPS. COVID-19: a chronology of state and territory government announcements (up until 30 June 2020). 2020;21.

37. Gray RT, Wilson DP, Guy RJ, Stoové M, Hellard ME, Prestage GP, et al. Undiagnosed HIV infections among gay and bisexual men increasingly contribute to new infections in Australia. Journal of the International AIDS Society. 2018 Apr;21(4):e25104.

38. Australian Bureau of Statistics. Overseas Migration. Canberra: ABS. 2022-23-financial-year [cited 2024 4th July]. Available from: https://www.abs.gov.au/statistics/people/population/overseas-migration/latest-release.

39. Cohen MS, Chen YQ, McCauley M, Gamble T, Hosseinipour MC, Kumarasamy N, et al. Prevention of HIV-1 infection with early antiretroviral therapy. The New England journal of medicine. 2011 Aug 11;365(6):493–505.

40. Rodger AJ, Cambiano V, Bruun T, Vernazza P, Collins S, Degen O, et al. Risk of HIV transmission through condomless sex in serodifferent gay couples with the HIV-positive partner taking suppressive antiretroviral therapy (PARTNER): final results of a multicentre, prospective, observational study. Lancet (London, England). 2019 Jun 15;393(10189):2428–38.

41. McCormack S, Dunn DT, Desai M, Dolling DI, Gafos M, Gilson R, et al. Pre-exposure prophylaxis to prevent the acquisition of HIV-1 infection (PROUD): effectiveness results from the pilot phase of a pragmatic open-label randomised trial. Lancet (London, England). 2016 Jan 2;387(10013):53–60.

42. Smith DK, Herbst JH, Zhang X, Rose CE. Condom effectiveness for HIV prevention by consistency of use among men who have sex with men in the United States. Journal of acquired immune deficiency syndromes (1999). 2015 Mar 1;68(3):337–44.

