## Supplementary Material for "Evaluating the impact of COVID-19 on the HIV epidemic among men who have sex with men in Australia: A modelling study"

Rongxing Weng^1§^, Jisoo A. Kwon^1^, Mo Hammoud^1^, Brent Clifton^2^, Nick Scott^3^, Skye McGregor^1^, and Richard T. Gray^1^

1 Kirby Institute, UNSW Sydney, Sydney, NSW, Australia;
2 National Association of People With HIV Australia, Sydney, NSW, Australia;

3	Burnet Institute, Melbourne, VIC, Australia

**Methodological details**

Additional methodological details for the data in the COVID-19 scenario and no COVID-19 plus PrEP scenario are provided as follows.

**COVID scenario: a COVID scenario with all changes in place**

To estimate the impact of the COVID-19 pandemic on the use of PrEP, we calculated the relative change in the monthly number of individuals dispensed PrEP, using data from the Kirby Institute’s project: Monitoring HIV pre-exposure prophylaxis (PrEP) uptake in Australia. We assumed that it could be used as an indicator of the relative change in the proportion of men who have sex with men (MSM) on PrEP, expressed as $\omega_{t}{/\omega}_{0}$. As the collection of the data was from January 2020, the number in December 2019 was considered the same as that in January 2020.

To estimate the impact of the COVID-19 pandemic on HIV testing, we used the monthly relative change in the proportion of HIV tested in the last four weeks as a proxy for the monthly relative change in the proportion of MSM tested $(T_{t}{/T}_{0})$. Data on the proportion of MSM tested for HIV within the last four weeks was collected by the Flux study in 2019, every four weeks from May 2020 to April 2021, and quarterly thereafter until August 2022. To adapt this variable to a monthly dataset, we first treated the data on a weekly basis. Linear regression was then applied to estimate missing data points between two available data points, followed by the aggregation of these weekly estimates into monthly averages. We assumed that the testing rates in January and February 2020 remained the same as pre-COVID-19 period and the rate in April 2020 was the same as that in May 2020. As Australia introduced physical distancing measures in mid-March 2020, we assumed that the impact of the pandemic on HIV testing for that month was half of that in April. The complete monthly data from December 2019 to August 2022 for $T_{t}{/T}_{0}$ was shown in Table S1.

To estimate the impact of the COVID-19 pandemic on sexual partners, we used the monthly number of non-romantic partners (casual sexual partners and friends with benefits) in the last week as a proxy for the monthly number of sexual partners $(p_{t})$. Data on the number of casual sexual partners and friends with benefits in the last week among MSM was collected by the Flux study weekly from May 2020 to April 2021, and quarterly thereafter until August 2022. To adapt this variable to a monthly dataset, we first treated the quarterly data on a weekly basis. Linear regression was then applied to estimate missing data points between two available data points, followed by the aggregation of these weekly estimates into monthly averages. We assumed that the number of sexual partners in January and February 2020 remained the same as pre-COVID-19 period. We used the ratio of the number of casual sexual partners between pre-COVID-19 and COVID-19 periods to calculate the value in this period. We assumed that the number of sexual partners in April 2020 was the same as that in May 2020. As Australia introduced physical distancing measures in mid-March 2020, we assumed that the impact of the pandemic on the number of sexual partners for that month was half of that in April. The complete monthly data from December 2019 to August 2022 for $p_{t}/p_{0}$ was shown in Table S1.

To estimate the impact of the COVID-19 pandemic on condom use, we used the monthly data on the proportion of MSM with casual partners reporting no condomless anal intercourse (CLAI) in the past four weeks as a proxy for the proportion of MSM reporting condom use at timestep *t* $(c_{t})$. Data on the proportion of MSM with casual partners reporting no CLAI in the past seven days was collected by the Flux study weekly from May 2020 to April 2021, and quarterly thereafter until August 2022. Data on the proportion of MSM with casual partners reporting no CLAI in the past six months was collected by the Gay Community Periodic Survey (GCPS) annually including the pre-COVID-19 period in February 2020. To convert these variables into a consistent monthly dataset, we assumed a weekly declining trend in the proportion of no CLAI, which follows the Poisson distribution. Using the data in 2022 from both the Flux study and GCPS, a Poisson regression model was fitted to explore the relationship between time and the proportion of no CLAI. In this regression, weekly timestep was considered as the dependent variable in which the no CLAI in the last week was set as the first timestep and no CLAI in the past six months was set as the 26^th^ timestep. Then, using this regression model, the pre-COVID-19 data on the proportion of no CLAI in the past six months from the GCPS was converted to the proportion of no CLAI in the past four weeks ($c_{0}$). Similarly, the weekly data on the proportion of no CLAI in the last week from the Flux study were converted to the proportion of no CLAI in the past four weeks, followed by the aggregation of the weekly estimates in the Flux study into monthly averages $(c_{t})$. We assumed that the proportion of no CLAI in the past four weeks in January and February 2020 remained the same as pre COVID-19 period. We assumed that the proportion of no CLAI in the past four weeks in April 2020 was the same as that in May 2020. As Australia introduced physical distancing measures in mid-March 2020, we assumed that the impact of the pandemic on the proportion of no CLAI for that month was half of that in April. The complete monthly data from December 2019 to August 2022 for the relative change of $c_{t}/c_{0}$ was shown in Table S1.

Given that the data on the proportion of diagnosed people with HIV taking antiretroviral therapy (ART) and the proportion of those taking ART with suppressed virus were only available on an annual basis, we first treated these annual values as the final month of each year. Linear regression was then applied to estimate missing data points between two available data points.

**No COVID-19 plus PrEP scenario: a no COVID-19 scenario continued PrEP scale-up**

To estimate the increase in the proportion of MSM on PrEP, we used the relative change in the number of people dispensed PrEP in the past three months as a proxy. Data on the number of people dispensed PrEP in the past three months was collected quarterly by the Kirby Institute’s project: Monitoring HIV pre-exposure prophylaxis (PrEP) uptake in Australia. To adapt this variable to a monthly dataset, we first treated the quarterly data on a monthly basis. A steady trend was found between February 2019 and February 2020, so we used them to fit a linear regression model, which was then applied to estimate data points from January 2020 to August 2022. To ensure consistency, the number in December 2019 was considered the same as that in January 2020. The complete monthly data from December 2019 to August 2022 for $\omega_{t}{/\omega}_{0}$ was shown in Table S1.

To estimate the decrease in the proportion of condom use, we used the monthly relative change in the proportion of MSM with casual partners reporting no CLAI in the past four weeks as a proxy. Using the previously fitted Poisson regression model, data on the proportion of no CLAI in the past six months from the GCPS in 2018, 2019, and 2020 was converted to the proportion of no CLAI in the past four weeks. A linear regression model was then fitted and applied to estimate data points from January 2020 to August 2022. The complete monthly data from December 2019 to August 2022 for the relative change of $c_{t}/c_{0}$ was shown in Table S1.

Table S1. Relative changes of four variables per month in COVID scenario and no COVID-19 plus PrEP scenario and the monthly number of MSM who enter Australia living with HIV in the COVID scenario

| **Time** | **Sexual partners (S1)** | **Condom** | **Use** | **HIV Testing (S1)** | **PrEP** | **use** | **Immigrants**  **(S1)** |
| --- | --- | --- | --- | --- | --- | --- | --- |
|  |  | **S1** | **S3** |  | **S1** | **S3** |  |
| Jan 2020 | 1.00 | 1.00 | 1.00 | 1.00 | 1.00 | 1.00 | 18 |
| Feb 2020 | 1.00 | 1.00 | 0.99 | 1.00 | 0.99 | 1.02 | 18 |
| Mar 2020 | 0.67 | 1.04 | 0.99 | 0.79 | 1.06 | 1.04 | 18 |
| Apr 2020 | 0.35 | 1.08 | 0.99 | 0.58 | 0.63 | 1.06 | 15 |
| May 2020 | 0.35 | 1.08 | 0.98 | 0.58 | 0.79 | 1.08 | 15 |
| Jun 2020 | 0.43 | 1.06 | 0.98 | 0.64 | 0.88 | 1.10 | 15 |
| Jul 2020 | 0.40 | 1.05 | 0.98 | 0.79 | 0.91 | 1.12 | 13 |
| Aug 2020 | 0.43 | 1.06 | 0.97 | 0.81 | 0.85 | 1.13 | 13 |
| Sep 2020 | 0.47 | 1.03 | 0.97 | 0.75 | 0.88 | 1.15 | 13 |
| Oct 2020 | 0.45 | 1.03 | 0.97 | 0.81 | 0.92 | 1.17 | 10 |
| Nov 2020 | 0.53 | 0.99 | 0.96 | 0.91 | 0.93 | 1.19 | 10 |
| Dec 2020 | 0.56 | 0.99 | 0.96 | 0.97 | 1.01 | 1.21 | 10 |
| Jan 2021 | 0.67 | 0.98 | 0.95 | 0.78 | 0.99 | 1.23 | 6 |
| Feb 2021 | 0.56 | 1.01 | 0.95 | 0.94 | 0.97 | 1.25 | 6 |
| Mar 2021 | 0.62 | 0.98 | 0.95 | 0.94 | 1.07 | 1.27 | 6 |
| Apr 2021 | 0.66 | 0.99 | 0.94 | 0.86 | 0.95 | 1.29 | 7 |
| May 2021 | 0.82 | 1.02 | 0.94 | 0.85 | 1.02 | 1.31 | 7 |
| Jun 2021 | 1.15 | 1.02 | 0.94 | 0.96 | 1.00 | 1.33 | 7 |
| Jul 2021 | 1.10 | 1.04 | 0.93 | 0.93 | 0.94 | 1.35 | 7 |
| Aug 2021 | 0.95 | 1.07 | 0.93 | 0.86 | 0.87 | 1.36 | 7 |
| Sep 2021 | 0.82 | 1.10 | 0.93 | 0.79 | 0.83 | 1.38 | 7 |
| Oct 2021 | 0.98 | 1.07 | 0.92 | 0.81 | 0.92 | 1.40 | 8 |
| Nov 2021 | 1.22 | 1.01 | 0.92 | 0.85 | 0.99 | 1.42 | 8 |
| Dec 2021 | 1.36 | 0.97 | 0.92 | 0.90 | 1.05 | 1.44 | 8 |
| Jan 2022 | 1.19 | 0.96 | 0.91 | 0.93 | 0.96 | 1.46 | 12 |
| Feb 2022 | 1.02 | 0.96 | 0.91 | 0.95 | 0.97 | 1.48 | 12 |
| Mar 2022 | 0.91 | 0.96 | 0.91 | 0.96 | 1.05 | 1.50 | 12 |
| Apr 2022 | 0.91 | 0.97 | 0.90 | 0.93 | 0.98 | 1.52 | 14 |
| May 2022 | 0.91 | 0.97 | 0.90 | 0.90 | 1.05 | 1.54 | 14 |
| Jun 2022 | 0.91 | 0.98 | 0.90 | 0.89 | 1.02 | 1.56 | 14 |
| Jul 2022 | 0.91 | 0.97 | 0.89 | 0.90 | 1.05 | 1.58 | 17 |
| Aug 2022 | 0.91 | 0.96 | 0.89 | 0.91 | 1.07 | 1.60 | 17 |

Numbers for relative change were rounded to two decimal places. Numbers for immigrants were rounded to an integer.

Abbreviations: HIV, human immunodeficiency virus; MSM, men who have sex with men; PrEP, preexposure prophylaxis.

*S1: COVID-19 scenario. †S3: No COVID-19 plus PrEP scenario

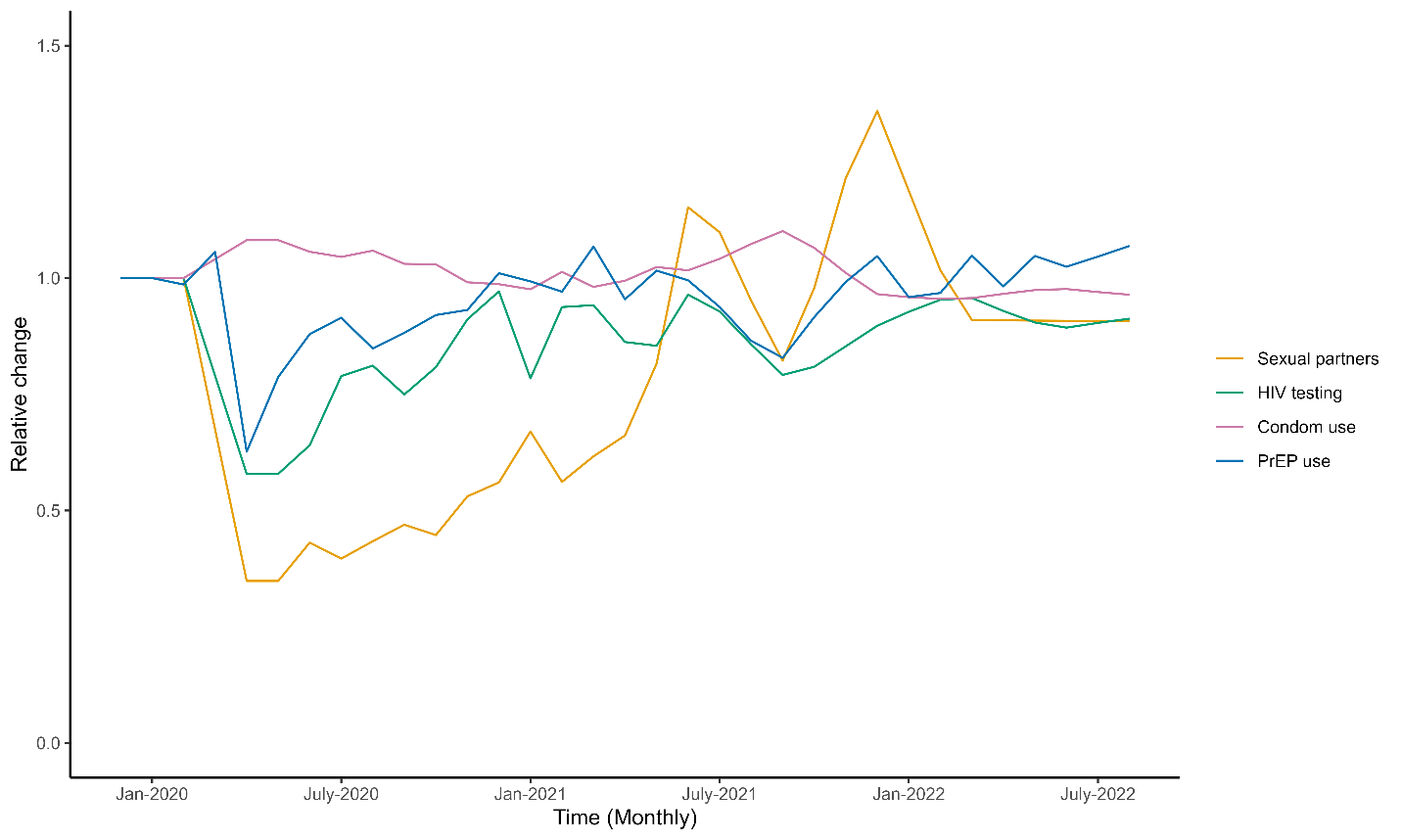

**Figure S1. Relative change of each time series variable compared to that in pre COVID-19 period.**

**Additional Results**

Additional details for the results are provided as the following.

Table S2. Monthly number of new HIV infections with 95% percentile intervals in each scenario

| **Time** | **New infections (S1*****)** | **Lower bound (S1)** | **Upper bound (S1)** | **New infections (S2**†**)** | **Lower bound (S2)** | **Upper bound (S2)** | **New infections (S3**‡**)** | **Lower bound (S3)** | **Upper bound (S3)** |
| --- | --- | --- | --- | --- | --- | --- | --- | --- | --- |
| Jan 2020 | 42 | 34 | 52 | 42 | 37 | 47 | 42 | 37 | 47 |
| Feb 2020 | 42 | 34 | 53 | 42 | 37 | 47 | 42 | 37 | 47 |
| Mar 2020 | 26 | 21 | 33 | 41 | 37 | 46 | 41 | 37 | 46 |
| Apr 2020 | 15 | 12 | 20 | 41 | 37 | 46 | 41 | 36 | 46 |
| May 2020 | 14 | 12 | 18 | 41 | 36 | 46 | 40 | 36 | 45 |
| Jun 2020 | 18 | 14 | 22 | 41 | 36 | 46 | 40 | 35 | 45 |
| Jul 2020 | 16 | 13 | 20 | 41 | 36 | 46 | 40 | 35 | 45 |
| Aug 2020 | 18 | 14 | 22 | 41 | 36 | 46 | 39 | 34 | 44 |
| Sep 2020 | 19 | 16 | 24 | 40 | 36 | 46 | 39 | 34 | 44 |
| Oct 2020 | 18 | 15 | 23 | 40 | 35 | 46 | 38 | 34 | 44 |
| Nov 2020 | 22 | 18 | 28 | 40 | 35 | 46 | 38 | 33 | 43 |
| Dec 2020 | 22 | 18 | 28 | 40 | 35 | 46 | 38 | 33 | 43 |
| Jan 2021 | 27 | 22 | 34 | 40 | 35 | 46 | 37 | 32 | 43 |
| Feb 2021 | 22 | 18 | 27 | 40 | 35 | 46 | 37 | 32 | 43 |
| Mar 2021 | 23 | 19 | 29 | 40 | 34 | 46 | 36 | 32 | 42 |
| Apr 2021 | 25 | 21 | 32 | 39 | 34 | 46 | 36 | 31 | 42 |
| May 2021 | 29 | 23 | 37 | 39 | 34 | 46 | 36 | 31 | 42 |
| Jun 2021 | 41 | 33 | 53 | 39 | 34 | 46 | 35 | 30 | 42 |
| Jul 2021 | 39 | 31 | 50 | 39 | 34 | 46 | 35 | 30 | 42 |
| Aug 2021 | 33 | 26 | 43 | 39 | 33 | 46 | 35 | 29 | 41 |
| Sep 2021 | 28 | 22 | 36 | 39 | 33 | 46 | 34 | 29 | 41 |
| Oct 2021 | 33 | 26 | 43 | 39 | 33 | 47 | 34 | 28 | 41 |
| Nov 2021 | 41 | 32 | 54 | 39 | 33 | 47 | 34 | 28 | 41 |
| Dec 2021 | 46 | 36 | 62 | 39 | 32 | 47 | 33 | 28 | 40 |
| Jan 2022 | 42 | 32 | 57 | 39 | 32 | 47 | 33 | 27 | 40 |
| Feb 2022 | 36 | 27 | 49 | 38 | 32 | 47 | 32 | 27 | 40 |
| Mar 2022 | 31 | 23 | 42 | 38 | 32 | 47 | 32 | 26 | 40 |
| Apr 2022 | 31 | 23 | 42 | 38 | 32 | 47 | 32 | 26 | 39 |
| May 2022 | 29 | 22 | 41 | 38 | 31 | 47 | 31 | 26 | 39 |
| Jun 2022 | 29 | 22 | 41 | 38 | 31 | 48 | 31 | 25 | 39 |
| Jul 2022 | 29 | 21 | 41 | 38 | 31 | 48 | 31 | 25 | 39 |
| Aug 2022 | 28 | 20 | 40 | 38 | 31 | 48 | 30 | 24 | 39 |

Numbers were rounded to an integer.

Abbreviations: HIV, human immunodeficiency virus.

*S1: COVID-19 scenario. †S2: No COVID-19 scenario. ‡S3: No COVID-19 plus PrEP scenario.

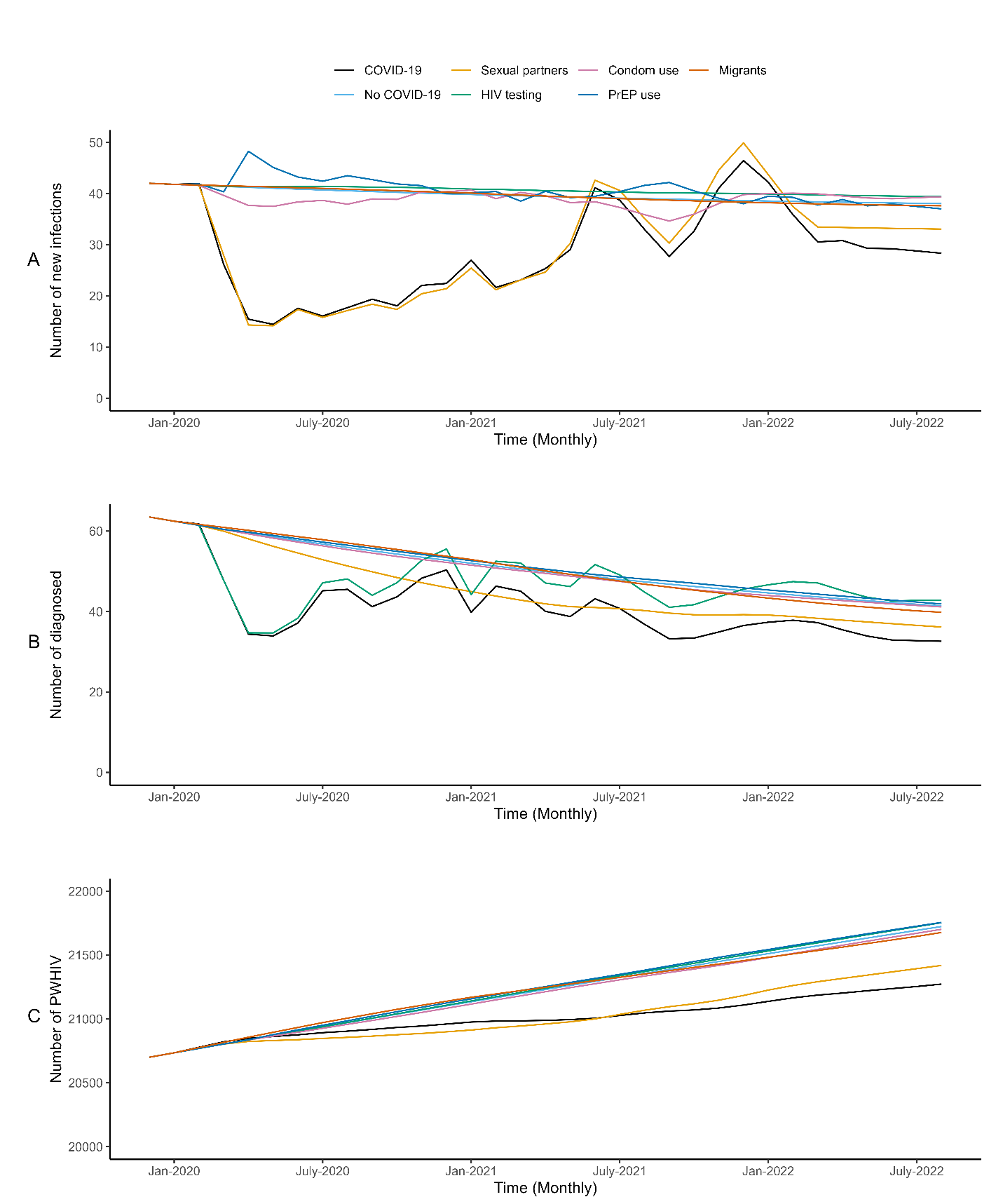

Figure S2. Impacts of each time series variable on HIV epidemic. (A) Monthly number of new HIV infections in COVID-19 scenario, no COVID-19 scenario, and scenarios including COVID-19 impact on each variable alone. (B) Monthly number of diagnoses in COVID-19 scenario, no COVID-19 scenario, and scenarios including COVID-19 impact on each variable alone. (C) Monthly number of people with HIV in COVID-19 scenario, no COVID-19 scenario, and scenarios including COVID-19 impact on each variable alone.

Table S3. Number of cumulative infections, cumulative diagnoses, and MSM living with HIV in no COVID-19 scenario and scenarios including COVID-19 impact on each variable alone between January 2020 to August 2022

| **Variables** | **Value** | **Relative change** |
| --- | --- | --- |
| Cumulative infections |  |  |
| No COVID-19 | 1,266 | Reference |
| Only include COVID-19 impact on sexual partners | 952 | -24.8% |
| Only include COVID-19 impact on HIV testing | 1,298 | 2.5% |
| Only include COVID-19 impact on condom use | 1,245 | -1.7% |
| Only include COVID-19 impact on PrEP use | 1,298 | 2.5% |
| Only include COVID-19 impact on migrants | 1,265 | -0.1% |
| Cumulative diagnoses |  |  |
| No COVID-19 | 1,613 | Reference |
| Only include COVID-19 impact on sexual partners | 1,443 | -10.5% |
| Only include COVID-19 impact on HIV testing | 1,492 | -7.5% |
| Only include COVID-19 impact on condom use | 1,599 | -0.8% |
| Only include COVID-19 impact on PrEP use | 1,631 | 1.1% |
| Only include COVID-19 impact on migrants | 1,608 | -0.3% |
| MSM living with HIV |  |  |
| No COVID-19 | 21,724 | Reference |
| Only include COVID-19 impact on sexual partners | 21,419 | -1.4% |
| Only include COVID-19 impact on HIV testing | 21,754 | 0.1% |
| Only include COVID-19 impact on condom use | 21,702 | -0.1% |
| Only include COVID-19 impact on PrEP use | 21,756 | 0.1% |
| Only include COVID-19 impact on migrants | 21,677 | -0.2% |

Abbreviations: HIV, human immunodeficiency virus; MSM, men who have sex with men; PrEP, preexposure prophylaxis.
